# Diagnostic accuracy of point-of-care lung ultrasound for COVID-19: A systematic review and meta-analysis

**DOI:** 10.1101/2021.10.09.21264799

**Authors:** Ashley K. Matthies, Michael M. Trauer, Karl Chopra, Robert Jarman

## Abstract

**Background:** Point-of-care (POC) lung ultrasound (LUS) is widely used in the emergency setting and there is an established evidence base across a range of respiratory diseases, including previous viral epidemics. The necessity for rapid testing combined with the limitations of other diagnostic tests has led to the proposal of various potential roles for LUS during the COVID-19 pandemic. This systematic review and meta-analysis focused specifically on the diagnostic accuracy of LUS in adult patients presenting with suspected COVID-19.

**Methods:** Traditional and grey-literature searches were performed on June 1^st^ 2021. Two authors independently carried out the searches, selected studies and completed the Quality Assessment Tool for Diagnostic Test Accuracy Studies (QUADAS-2). Meta-analysis was carried out using established open-source packages in *R*. We report overall sensitivity, specificity, positive and negative predictive values and the hierarchical summary receiver operating characteristic curve for LUS. Heterogeneity was determined using the I^2^ statistic.

**Results:** Twenty studies were included, providing data from a total of 4,314 patients. The prevalence and admission rates were generally high across all studies. Overall LUS was found to be 87.2% sensitive (95% CI 83.6-90.2) and 69.5% specific (95% CI 62.2-72.5) and demonstrated overall positive and negative predictive values of 3.0 (95% 2.3-4.1) and 0.16 (95% 0.12-0.22) respectively. Separate analyses for each reference standard revealed similar sensitivities and specificities for LUS. Heterogeneity between studies was found to be high, and QUADAS-2 assessment identified risks of bias in many studies.

**Conclusion:** During a period of high prevalence, LUS is a highly sensitive diagnostic test for COVID-19. However, more research is required to confirm these results in more generalisable populations, including those less likely to be admitted to hospital.

## Introduction

Coronavirus disease 2019 (COVID-19) was declared a global pandemic by the World Health organization on the 11th March 2020. As of the 27th August 2021 there have been more than 214 million confirmed cases and more than 4.4 million deaths.(1)

There is now an established evidence base concerning the role of point-of-care (POC) lung ultrasound (LUS) in patients presenting with acute respiratory distress.(2) First described in 2008, the BLUE (Bedside Lung Ultrasound in Emergency) protocol was found to be greater than 90% accurate for the underlying diagnosis of acute respiratory distress. Since then, there has been a proliferation of research into LUS which has confirmed that it is highly accurate for several respiratory conditions and often more accurate than chest radiograph (CXR).(3, 4)

The utility of LUS has also been described during previous viral epidemics. LUS was found to be accurate in differentiating viral and bacterial pneumonia during the influenza epidemic in 2009 and was found to be more accurate than CXR for avian flu.(5, 6)

Furthermore, CXR and a single initial reverse transcriptase polymerase chain reaction (RT-PCR) test have both been shown to have sub-optimal diagnostic accuracy for COVID-19. In a large meta-analysis, CXR was found to be 81% sensitive and 72% specific.(7) A single initial RT-PCR test has also been found to have a sensitivity as low as 71% when compared to serial testing.(8) Whilst chest Computed Tomography (CT) has been shown to be more accurate than CXR (7), this is neither an appropriate nor feasible test to provide for every patient, and is not recommended by the Royal College of Radiologists.(9)

As a result various roles for LUS have been proposed including triage, diagnosis, prognostication and monitoring of disease progression.(10) Multiple studies have demonstrated that LUS may have superior test characteristics than CXR.(11-13) Furthermore, LUS is a rapid test that can be performed at the bedside, causes no radiation exposure and has good inter-operator characteristics.(14) It may also have the potential to reduce nosocomial infections and healthcare provider exposure to COVID-19.(15)

The sonographic findings of COVID-19 pneumonitis (or pneumonia) are now well described and include pleural line abnormalities, B-lines and consolidation. The constellation of findings is also well understood with abnormalities typically bilateral and patchy.(16)

A scoping review of the diagnostic utility of LUS in COVID-19 was published in August 2020, yielding 33 studies. The methodology of the included studies was generally poor and only 5 studies reported diagnostic accuracy. However, there was a trend towards high sensitivity and low specificity.(17)

Since then, we have experienced second and third waves of infection, and further evidence concerning the role of LUS has accumulated. The purpose of this systematic review was to evaluate the diagnostic accuracy of POC LUS in adult patients presenting with suspected COVID-19 infection compared to three commonly used reference standards used for the diagnosis of COVID-19; 1) RT-PCR, 2) chest CT, 3) Aggregate final clinical diagnosis.

## Methods

This systematic review was synthesised according to the Preferred Reporting Items for Systematic Reviews and Meta-analyses(18), and the protocol was registered on PROSPERO (CRD42021250464). No funding was sought for the production of this work.

### Search strategy

Traditional sources of literature were searched, including Ovid MEDLINE, Embase, SCOPUS, Cochrane Library and Google Scholar. Less traditional sources were also searched including medRxiv, the pre-print server for health sciences.

A search strategy was developed in line with existing guidance(19) and searches were performed on June 1^st^ 2021 (see supplementary material 1).

### Eligibility

All prospective and retrospective trials of patients over 16 years of age comparing lung ultrasonography to either RT-PCR, chest CT, or a final clinical diagnosis of COVID-19 were included. Case reports, editorials and recommendation/instructional journal papers were excluded.

### Study selection

Searches were carried out independently by two reviewers (AM, MT), with disagreements reviewed by a third reviewer (RJ). A screening and selection tool (see supplementary material 2) was agreed upon and applied to identify eligible studies from both the abstract and subsequent full-text review. The reference lists from these studies were also reviewed to identify and further relevant studies.

### Definition of the index test / diagnostic threshold

Multiple scanning protocols have been described and the specific threshold of findings necessary to diagnose COVID-19 has also not been agreed upon. Studies were included regardless of the number of zones scanned or the diagnostic threshold used.

### Definition of the reference test

Although a positive RT-PCR is recognised as the Gold Standard diagnostic test for ruling-in COVID-19 infection, it is less reliable at ruling-out the disease. Whilst RT-PCR testing is used to detect infection LUS is used to diagnose viral pneumonia. Chest CT has been described as the reference standard for viral pneumonia(20) but is infrequently used in clinical practice. An alternative to both is the use of a final clinical diagnosis incorporating imaging findings and results of serial RT-PCR tests. We have included studies using all three reference standards, and present combined and individual data.

### Data collection and synthesis

Two independent reviewers (AM, MT) extracted the following data (displayed in Table 1): Study design, exclusion criteria, number of patients, admission rate (used as a measure of disease severity), prevalence of COVID-19, sensitivity, specificity, number of true positive and negative results, number of false positive and negative results, scanning technique and diagnostic threshold (or scoring) for LUS, reference test and blinding. Disagreements were resolved by a third independent reviewer (RJ).

**Table 1.** Summary of key demographic and epidemiological data as well as results from each of the included studies, arranged by sample size from highest.

| Author | Submission (publication) date | Design / enrolment | Exclusion criteria | n | COVID-19 prevalence (%) | Admission rate (%) | LUS protocol | LUS Dx threshold | Reference test | Sensitivity LUS | Specificity LUS | TP | FN | FP | TN | Prior training and experience |
| --- | --- | --- | --- | --- | --- | --- | --- | --- | --- | --- | --- | --- | --- | --- | --- | --- |
| **Volpicelli *et al*** | December 2020 (March 2021) | Prospective, convenience | None | 1462 | 70 | 34 | 'Complete' ($\geq 12$ zones) | High or Intermediate pattern | PCR | 90 | 53 | 921 | 101 | 209 | 231 | Documented evidence in performing LUS |
| **Colombi *et al*** | July 2020 (October 2020) | Retrospective, convenience | Those without clinic or epidemiological suspicion | 486 | 70 | 100 | 12 zones | $\geq 1$ zone | PCR | 91 | 66 | 311 | 29 | 49 | 97 | No details provided |
| **Sorlini *et al*** | June 2020 (October 2020) | Retrospective, convenience | None | 384 | - | 100 | 12 zones | $\geq 2$ zones positive | PCR | 92 | 65 | 264 | 23 | 34 | 63 | LUS accreditation, 1-day LUS course and >150 supervised scans |
| **Bianchi *et al*** | October 2020 (April 2021) | Prospective, convenience | Pregnancy, confirmed COVID-19 infection on PCR | 360 | 39 | 100 | 12 zones | Summary assessment | PCR | 86 | 71 | 120 | 20 | 64 | 156 | LUS accreditation and >50 LUS examinations |
| **Pivetta *et al*** | July 2020 (October 2020) | Prospective, convenience | Intubated patients<br>Confirmed cases<br>Those requiring acute mental health care | 228 | 47 | 40 | 12 zones | Operator interpretation | PCR | 94 | 95 | 101 | 6 | 6 | 115 | LUS accreditation, Single learning module and >40 supervised scans |
| **Lieveld *et al*** | July 2020 (October 2020) | Prospective, convenience | Patients with uninterpretable LUS images | 187 | 46 | 100 | 12 zones | $\geq 2$ zones positive | PCR | 92 / 88 | 71 / 82 | 79 | 7 | 29 | 71 | Certified and >20 supervised scans |
| **Brenner *et al*** | December 2020 (March 2021) | Retrospective, convenience | None | 174 | 53 | 77 | 12 zones | Summary assessment | PCR | 86 | 72 | 79 | 13 | 23 | 59 | Credentialed in LUS |
| **Schmid *et al*** | July 2020 (December 2020) | Retrospective, convenience | None | 135 | 29 | 73 | 12 zones | $\geq 2$ zones positive | PCR / CT | 77 / 65 | 77 / 73 | 30 / 13 (PCR/CT) | 9 / 7 | 22 / 3 | 74 / 8 | Training in POCUS |
| **Zanforli**<br>**n et al** | June 2020<br>(December<br>2020) | Retrospective,<br>convenience | None | 111 | 41 | 48 | 20 zones | LUS score $\geq 3$ | Final Dx | 90 | 75 | 39 | 7 | 16 | 49 | No details provided |
| **Gibbons**<br>**et al** | November<br>2020<br>(February<br>2021) | Prospective,<br>convenience | Pregnancy | 110 | 75 | 100 | 8 zones | $\geq 2$ zones<br>positive | CT | 98 | 33 | 79 | 2 | 19 | 10 | ACEP credentialed<br>and >25 LUS scans |
| **Tung-**<br>**Chen et**<br>**al** | October 2020<br>(January<br>2021) | Prospective,<br>convenience | None | 96 | 61 | 77 | 12 zones | $\geq 2$ zones<br>positive | PCR | 72<br>(irregular<br>pleural line)<br>87<br>(confluent<br>b-lines) | 65<br>(irregular<br>pleural line)<br>33<br>(confluent<br>b-lines) | 39 | 15 | 12 | 23 | Experienced<br>sinologists,<br>performing >10 US<br>scans per week |
| **Haak et**<br>**al** | May 2020<br>(November<br>2020) | Prospective,<br>convenience | Age < 16<br>CCF<br>No LUS<br>Confirmed COVID | 93 | 29 | - | 12 zones | $\geq 1$ zone | PCR / CT | 96 / 89 | 59 / 59 | 23 / 3<br>(PCR<br>/ CT) | 1 / 2 | 28 / 8 | 41 /<br>13 | >2.5 years of<br>POCUS experience |
| **Narinx**<br>**et al** | June 2020<br>(September<br>2020) | Retrospective,<br>convenience | None | 90 | 17 | - | 12 zones | $\geq 1$ zone | PCR | 93 | 21 | 14 | 1 | 59 | 16 | Board certified for<br>LUS and >5 years<br>of experience in US |
| **Karagoz**<br>**et al** | May 2020<br>(December<br>2020) | Prospective,<br>convenience | Age < 18<br>Chest pain<br>Pregnant<br>Hypotensive<br>Previous thoracic surgery<br>If unable to obtain optimal<br>images | 72 | 48 | 100 | 12 zones | Not defined | CT | 92 | 97 | 32 | 1 | 3 | 36 | 5 years of<br>experience, 3-hour<br>theory module, 20<br>live demonstrations |
| **Jalil et**<br>**al** | September<br>2020 (October<br>2020) | Retrospective,<br>convenience | None | 69 | 52 | 100 | 8 zones | LUS-COV<br>criteria<br>(summary<br>assessment of<br>images) | PCR | 91 | 86 | 31 | 5 | 3 | 30 | No details<br>provided, but all 3<br>operators were<br>Intensive Care<br>physicians |
| **Fonsi et**<br>**al** | May 2020<br>(September<br>2020) | Prospective,<br>convenience | None | 63 | 70 | 100 | 12 zones | Not defined | PCR | 68 | 79 | 30 | 14 | 4 | 15 | No details provided |
| **Bosso**<br>**et al** | May 2020<br>(October<br>2020) | Prospective,<br>convenience | None | 53 | 49 | 100 | 12 zones | Severity score<br>used | PCR | 73 (optimal<br>cut-off<br>$\geq 12.5$) | 89 (optimal<br>cut-off<br>$\geq 12.5$) | 19 | 7 | 3 | 24 | No details provided |
| **Spiedel et al** | September 2020<br>(December 2020) | Prospective, convenience | If PCR result already available<br>ICU admissions | 49 | 22 | 100 | 12 zones | Severity score used | Final Dx | 91 (optimal cut-off score $\geq 8$) | 76 (optimal cut-off score $\geq 8$) | 10 | 1 | 9 | 29 | Specific POCUS qualification, >3 years of experience in US, >1 year experience of experience in LUS |
| **Walsh et al** | July 2020<br>(September 2020) | Retrospective, convenience | History of CCF or other pathology likely to affect LUS | 49 | 12 | 39 | Continuous posterior assessment + single anterior and axillary zones (equivalent to an 8-zone protocol) | Operator interpretation | CT | 100 | 88 | 12 | 0 | 2 | 8 | Facile in LUS, but no specific details provided |
| **Pare et al** | April 2020<br>(June 2020) | Retrospective, convenience | If there was no CXR, LUS or PCR test<br>If the patient had an underlying condition predisposing them to B-lines on LUS | 43 | 70 | 72 | Unclear (median number of saved images = 6) | $\geq 1$ B-line | PCR | 89 | 56 | 24 | 3 | 7 | 9 | No details provided. Images over-interpreted by 3 fellowship trained physicians. |

### Quality appraisal

Two reviewers (AM, MT) independently used the Quality Assessment Tool for Diagnostic Test Accuracy Studies (QUADAS-2) to evaluate the quality of each study and risk of bias. Disagreements were resolved by a third independent reviewer (RJ).

### Data analysis

Data was analysed using a dedicated application, MetaDTA (Complex Reviews Support Unit, National Institute for Health research, United Kingdom), a validated tool for meta-analysis of diagnostic test accuracy studies that utilises open-source packages in R (R Foundation for Statistical Computing, Vienna, Austria).(21) Random effects bivariate binomial models were used to estimate overall sensitivity, specificity and provide the hierarchical summary receiver operating characteristic (HSROC) curve for LUS. The open-source package *Metafor* in R was used to determine heterogeneity for univariate sensitivity and specificity analysis using a DerSimonian and Laird random effects model, providing the I^2^ statistic.(22, 23) Combined and separate analyses were performed for each of the reference standards.

## Results

Our search identified 3041 studies from traditional databases and registers, and 221 from other sources. Following removal of duplicates, eligibility screening by title/abstract and then full-text review a total of 20 studies were included. A flow chart was produced in line with the PRISMA guidelines (Figure 2).

**Figure 1.**
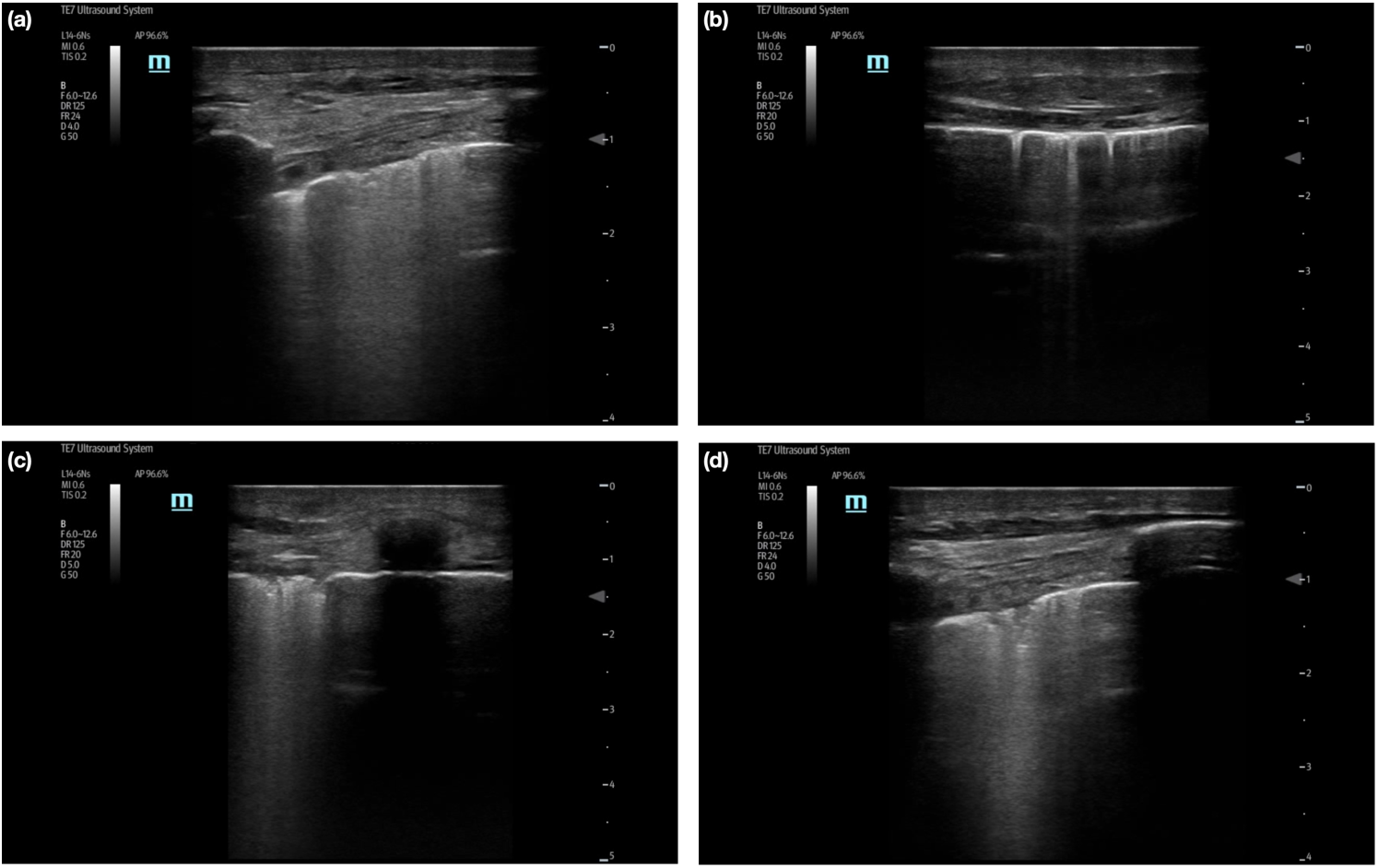
Typical LUS findings in COVID-19 pneumonia; (a) pleural line irregularity and confluent B-line formation, (b) only B-line formation, and in (c) and (d) peripheral (sub-pleural) consolidations, again with B-line formation.

**Figure 2.**
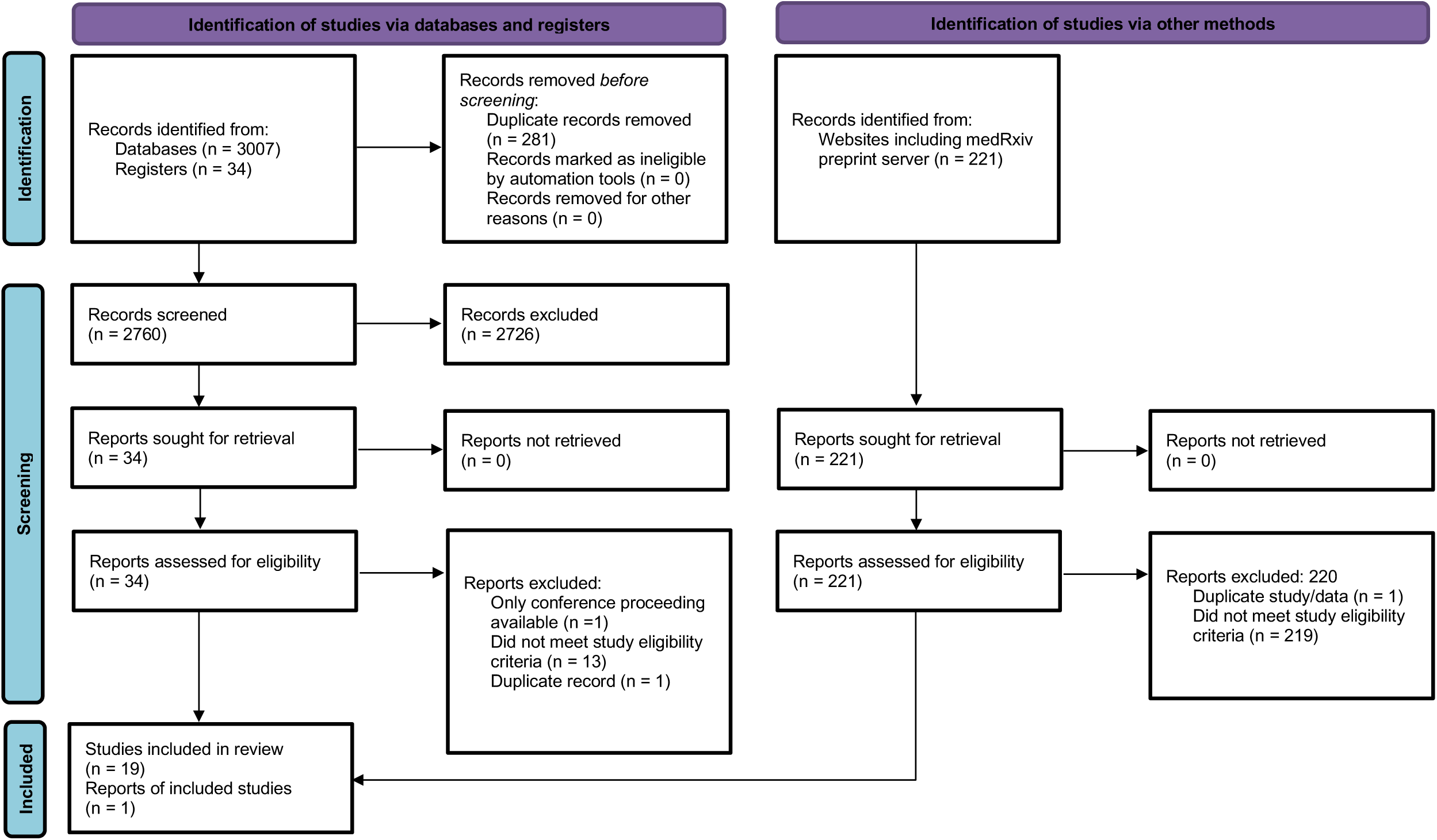
Prisma flow diagram

### Summary of results

An overview of the characteristics of all the included studies is shown in Table 1. A total of 4,314 patients were included. There was an even mix of prospective (n = 11) and retrospective (n = 9) studies, and all were convenience samples. The prevalence of COVID-19 infection ranged from 12% to 75%. Thirteen studies used RT-PCR as the reference test, whilst five used chest CT and three the final clinical diagnosis. The smallest study included 43 patients whilst the largest included 1462. Sensitivity of LUS ranged between 68% and 100% and specificity ranged between 21% and 97%. (11-14, 24-39)

### Comparison to RT-PCR testing

Fifteen studies used RT-PCR as the reference standard. In this group, sensitivity ranged between 68% and 96% whilst specificity ranged between 21% and 91%. (11, 12, 14, 24-35)

This included the largest study published study which included 1462 patients, and classified patients according to their clinical phenotype: 1) Mild (without dyspnoea or respiratory failure); 2) Severe (with dyspnoea or respiratory failure); and 3) Mixed (patients with cardiorespiratory comorbidities). LUS studies were classified as either low, intermediate or high probability of COVID, or suggestive of an alternate diagnosis. Interestingly, a high probability LUS demonstrated a modest sensitivity of 60.3% but a relatively strong specificity of 88.9, whilst combined high/intermediate LUS studies predictably demonstrated a higher sensitivity of 90.2% but lower specificity of 52.5%. In patients with an ‘alternate’ diagnosis (or disease pattern) on LUS, the majority tested negative on RT-PCR.

Three studies compared both LUS and chest CT to the reference standard RT-PCR result. In each study LUS demonstrated sensitivity greater than 90%, whereas the sensitivity of chest CT ranged between 69% and 90%. Both LUS and CT were far less specific than RT-PCR. (28, 31, 32). Three studies also found LUS to be more sensitive than CXR. (11, 12, 35)

In one study, the authors compared traditional clinical evaluation to an integrated assessment incorporating POC LUS. Clinical examination was found to be 81% sensitive and 64% specific whereas integrated assessment with LUS was 94% sensitive and 95% specific.

Two studies adopted scoring systems for the LUS findings. Unsurprisingly those who tested positive on RT-PCR had higher scores (optimal cut-off scores are shown in Table 1).(25, 26). It was also demonstrated that complete (12-zone) studies were 5% more sensitive and 4% more specific than less thorough studies. (26)

### Comparison to chest CT

Five studies (a total of 282 patients) used chest CT as the reference standard. Sensitivity ranged between 65% and 97% and specificity between 59% and 97%.(13, 27, 34, 36, 37)

In one of these studies, LUS was also compared to CXR and found to be more sensitive and similarly specific.(36)

### Comparison to final clinical diagnosis

Three studies used a final clinical diagnosis as the reference standard (a total of 347 patients). Sensitivity of LUS ranged between 85% and 91% and specificity between 75% and 80%. (31, 38, 39)

Two studies adopted scoring systems for LUS and optimal cut-off scores demonstrating sensitivities above 90% and specificities above 75%. (39)

#### Meta-analysis

Combining all studies, LUS had an overall sensitivity of 87.2% (95% CI 83.6-90.2) and specificity of 69.5% (62.2-72.5). Forest plots for each are shown in Figure 3a. Positive and negative likelihood ratios were 3.04 (2.27-4.06) and 0.16 (0.12-0.22) respectively. The HSROC curve is shown in Figure 4. Heterogeneity between all studies was high, with I^2^ values of 71.6% (95% CI 58.7-91.5) and 86.8% (84.4-96.0) for sensitivity and specificity respectively.

**Figure 3.**
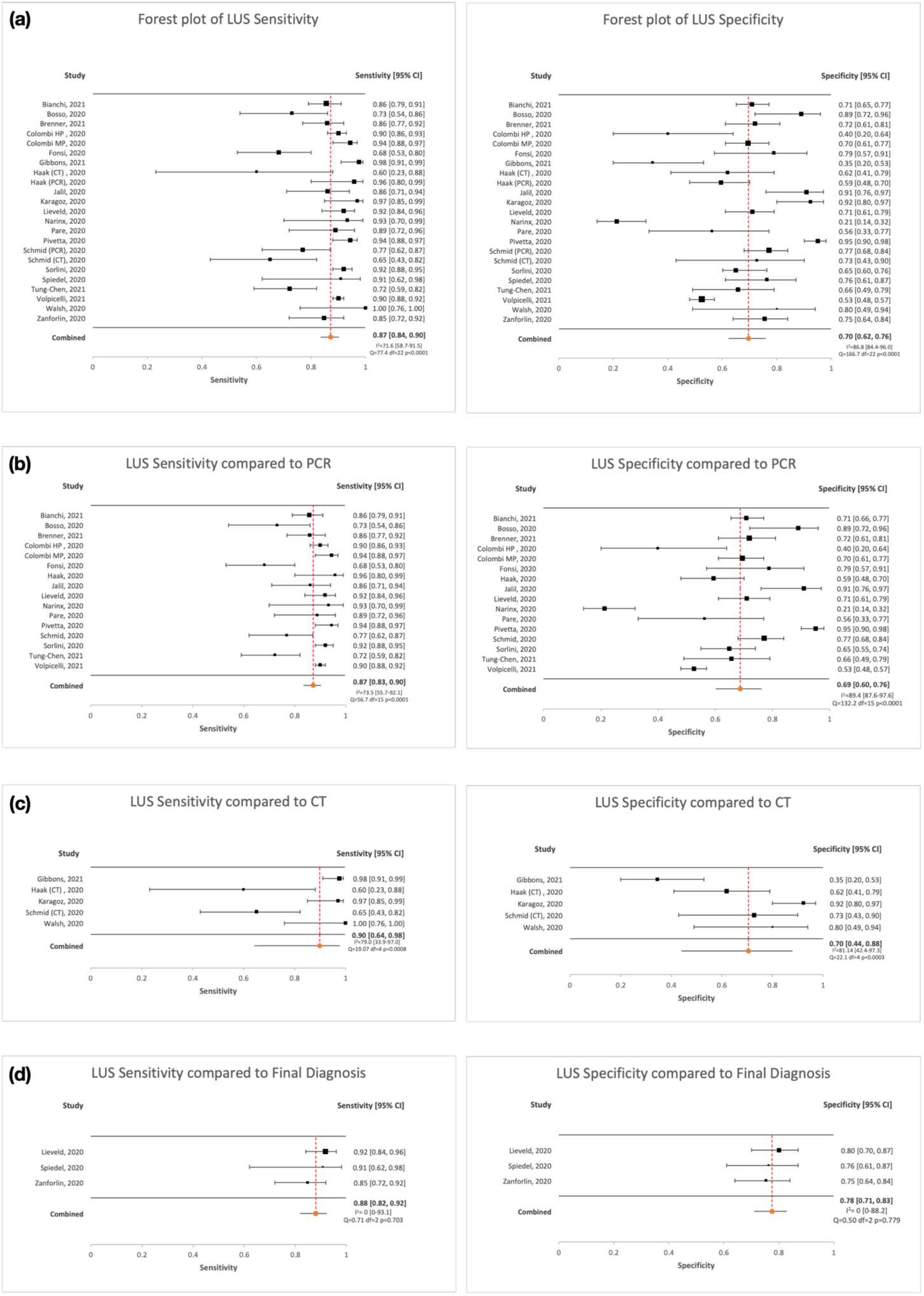
Forest plots demonstrating sensitivity and specificity of LUS overall (a) and compared to each of the three reference standards (RT-PCR (b), chest CT (c) and final clinical diagnosis (d))

**Figure 4.**
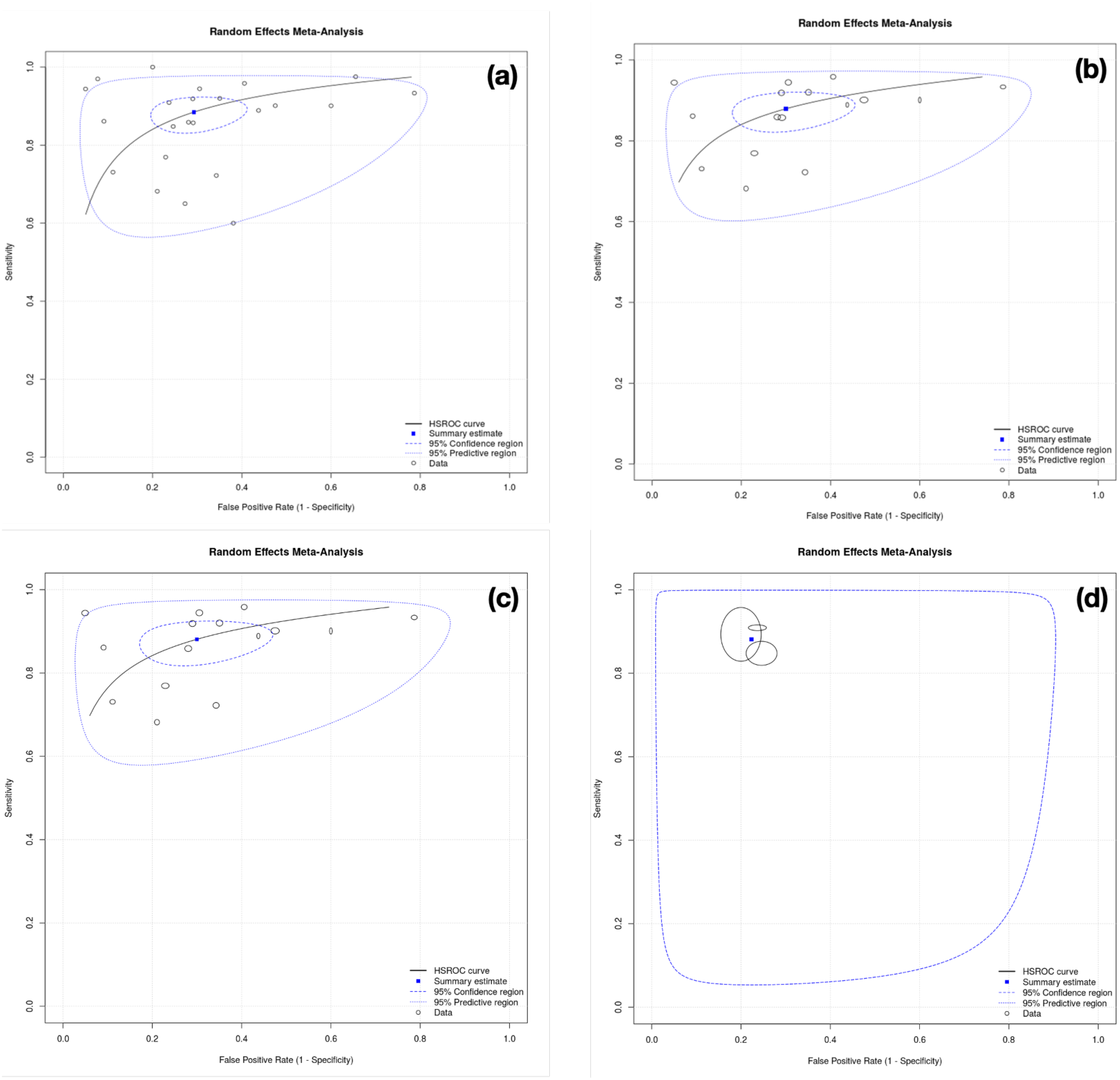
Hierarchical summary receiver operating characteristic (HSROC) curves for LUS overall (a) and compared to each of the three reference standards (RT-PCR (b), chest CT (c) and final clinical diagnosis (d))

When only studies using RT-PCR as the reference standard were included, sensitivity and specificity remained similar at 87.2% (83.4-90.3) and 68.7% (60.2-75.3) respectively. Forest plots are shown in Figure 3b. Positive and negative likelihood ratios were 2.94 (2.06-4.19) and 0.17 (0.12-0.24) respectively. Heterogeneity remained high with I^2^ values of 73.5% (55.7-92.1) and 89.4% (87.6-97.6) for sensitivity and specificity respectively.

When only the five studies using chest CT as the reference test were analysed, sensitivity increased to 89.7% (64.1-97.7) and specificity to 70.5% (43.9-87.9). Forest plots are shown in Figure 3c. Positive and negative likelihood ratios were 3.24 (1.56-6.73) and 0.10 (0.02-0.47) respectively. Again, heterogeneity was high, with I^2^ values of 79.0% (33.9-97.0) and 81.1% (42.4-97.3) for sensitivity and specificity respectively.

When only the studies comparing LUS to a final clinical diagnosis were examined, sensitivity remained similar at 88.0% (81.9-92.2) however specificity increased to 77.6% (71.1-83.0). Forest plots are shown in Figure 3d. Heterogeneity between these three studies was low with I^2^ values of 0% (0.0-88.1) and 0% (0.0-0.81).

HSROC curves for LUS compared to each of the reference standards are shown in Figures 4b-d.

#### Quality appraisal

The results of the QUADAS-2 assessment are shown in Table 2. Overall, most studies were deemed high risk for bias. Convenience sampling and the use of certain exclusion criteria introduced a risk of selection bias in all studies. There were also applicability concerns, with most studies conducted during a period of high disease prevalence and including predominantly patients requiring hospital admission.

**Table 2.** QUADAS-2 assessment (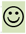 Low risk, 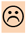 High risk, 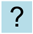 Unclear risk). Studies arranged by sample size, from largest.

| Study | Risk of Bias |  |  |  | Applicability concerns |  |  |
| --- | --- | --- | --- | --- | --- | --- | --- |
|  | Patient selection | Index test | Reference standard | Flow and timing | Patient selection | Index test | Reference standard |
| Volpicelli | 😞 | 😊 | 😊 | 😊 | 😞 | 😊 | 😊 |
| Colombi | 😞 | 😊 | 😊 | 😊 | 😞 | ? | 😊 |
| Sorlini | 😞 | 😊 | 😊 | 😊 | 😞 | 😊 | 😊 |
| Bianchi | 😞 | 😊 | 😊 | 😊 | 😞 | 😊 | 😊 |
| Pivetta | 😞 | 😊 | 😊 | 😊 | 😞 | 😊 | 😊 |
| Lieveld | 😞 | 😞 | 😊 | 😊 | 😞 | 😊 | 😊 |
| Brenner | 😞 | ? | 😊 | 😊 | 😞 | 😊 | 😊 |
| Schmid | 😞 | 😊 | 😊 | 😊 | 😞 | 😊 | 😊 |
| Zanforlin | 😞 | 😊 | 😞 | 😊 | 😞 | ? | 😞 |
| Gibbons | 😞 | ? | 😊 | 😊 | 😞 | 😊 | 😊 |
| Tung-Chen | 😞 | 😊 | 😊 | 😊 | 😞 | 😊 | 😊 |
| Haak | 😞 | 😊 | 😊 | 😊 | 😞 | 😊 | 😊 |
| Narinx | 😞 | 😊 | 😊 | 😊 | 😞 | 😊 | 😊 |
| Karagoz | 😞 | ? | 😊 | 😊 | 😞 | 😊 | 😊 |
| Jalil | 😞 | 😊 | 😊 | 😊 | 😞 | ? | 😊 |
| Fonsi | 😞 | ? | 😊 | 😊 | 😞 | 😊 | 😊 |
| Bosso | 😞 | 😊 | 😊 | 😊 | 😞 | 😊 | 😊 |
| Spiedel | 😞 | 😊 | 😞 | 😊 | 😞 | 😊 | 😞 |
| Walsh | 😞 | ? | 😊 | 😊 | 😞 | 😊 | 😊 |
| Pare | 😞 | 😞 | 😊 | 😊 | 😞 | 😊 | 😊 |

## Discussion

In this systematic review we aimed to evaluate the diagnostic accuracy of LUS for COVID-19 infection, diagnosed either by RT-PCR testing, chest CT scan or an aggregate final clinical diagnosis. We presented data from 20 studies consisting of more than 4,000 patients. The majority of studies were conducted in high prevalence populations with high rates of hospital admission. Overall, LUS was found to be highly sensitive but generally less specific for COVID-19 infection. The negative likelihood ratio (for ruling-out disease) was reasonable however the positive likelihood ratio was less useful. There was a high degree of variation between studies.

We reported test characteristics for LUS combining all studies and separately for the three reference standards commonly reported in the literature. However, it may be argued that in the population studied all three are in fact comparable. The rate of hospital admission was high across the studies and therefore it would be expected that the majority of patients were hypoxic with secondary complications of COVID-19 infection including pneumonia. (40) Given that chest CT is the recognised gold standard test for viral pneumonia,(20) it may therefore be considered broadly equivalent to RT-PCR testing in this population. This is supported by separate meta-analyses for each of the reference standards demonstrating similar pooled sensitivities (87%-90%) and specificities (69%-78%).

It is important to remember that LUS only tests for the pulmonary manifestations of COVID-19 and its sensitivity may therefore be significantly lower in non-admitted patients with mild or non-respiratory symptoms. The choice of reference test for comparison to LUS is therefore essential and we recommend future research examining LUS as a screening or diagnostic test for COVID-19 infection use RT-PCR testing, whereas studies examining the accuracy of LUS at diagnosing pulmonary manifestations should use chest CT.

Although in this study we reported sensitivity and specificity, examining the positive and negative predictive values (PPV and NPV), which incorporate the effect of disease prevalence can be useful measures of the performance of a diagnostic test. This is an important consideration given the overall high prevalence rates reported in the studies. In an era of relatively lower prevalence, the PPV of LUS may be significantly lower than has been estimated. Caution should therefore be taken when considering the application of LUS for early diagnosis, screening and cohorting patients in the setting of lower prevalence.

There is a rapidly expanding evidence base demonstrating favourable characteristics for molecular and other non-RT-PCR tests for COVID-19 that are much easier and faster to perform and can turn over rapid results.(8) The availability of these tests will likely impact on the value of LUS as an early bedside clinical tool.

Integrated LUS, used in combination with other clinical data, may also improve early diagnostic accuracy in the ED. Walsh *et al* showed that both sensitivity and specificity were increased when LUS was integrated with clinical findings as opposed to interpreted in isolation.(13) The potential to integrate bedside imaging findings is unique to POCUS and may enhance diagnostic accuracy. This concept was exemplified by Pivetta *et al* when they showed that an integrated LUS approach was highly accurate in patients with acute decompensated heart failure, more so than CXR or B-natriuretic peptide levels.(4)

All but three studies adopted a comprehensive 12-zone LUS technique. Given that the changes observed in viral pneumonia are bilateral and patchy, scanning technique is likely to affect sensitivity. Various scanning techniques have been described ranging from only limited views of the anterior chest to a more thorough examination of each intercostal space both anteriorly and posteriorly. Brenner *et al*, showed that when only complete 12-zone studies were included in their analysis, both sensitivity and specificity of LUS were improved.(26)

The heterogenous criteria used to determine whether a positive LUS may also have affected the results of the various. Various thresholds were used for ruling in this diagnosis, ranging from any B lines to a minimum number of affected zones to a minimum severity score.

Lower diagnostic thresholds generally favoured improved sensitivity whilst higher thresholds favoured specificity. Whilst relatively common in COVID-19 pneumonia, B-lines are also commonly seen in multiple respiratory diseases. (2) Intuitively, a low diagnostic threshold (for example, the presence of any B-lines) is likely to be sensitive but non-specific. Data presented by Volpicelli *et* al suggests that a diagnostic score weighted to the specific sonographic findings is likely to be more useful than a binary cut-off.(14) We therefore propose the development of a diagnostic (as opposed to severity) scoring system. Such a system could quantify the likelihood of COVID-19 depending on how typical the LUS features were for COVID-19. A resulting estimated likelihood ratio could then be integrated with a pre-test probability to provide the treating clinician with a post-test probability.

### Study limitations

The overall quality of studies was low, with internal validity affected by convenience sampling and the external validity by patient selection concerns. Furthermore, several studies reported data from small samples and provided no prior sample size estimates.

There was significant variation in the LUS protocols used as well as the diagnostic threshold for a positive scan. Furthermore, there was some variability regarding the prior experience and training of the operators. POCUS is an operator-dependent technique where a single provider is responsible for both image acquisition and interpretation. Therefore, both the specific training received and prior experience of the scanning physicians is likely to influence diagnostic accuracy.

### Future research

Future research should focus on wider populations, including mildly and asymptomatic patients, during periods of lower disease prevalence to define the role and setting in which LUS is most useful. As previously described, future work is required to define the optimal diagnostic threshold for LUS. Finally, further work is required to understand the training and experience required to gain proficiency in LUS.

### Recommendations / Conclusions

LUS was found to be highly sensitive in a population of both high prevalence and mostly admitted patients and may improve detection of COVID-19 pneumonia in this group compared to CXR. In hypoxic patients requiring admission, a normal LUS should prompt consideration of an alternative diagnosis. Although overall specificity was less optimal, the presence of a full spectrum or typical pattern of LUS findings is likely to be more specific and as such a quantitative diagnostic LUS scoring system is likely to be more useful than a binary threshold.

## Data Availability

All data produced in the present study are available upon reasonable request to the authors

## Supplementary material 1: Search terms and search strategy

Searches of traditional databases and registers were undertaken using the following key terms and strategy:

1. ‘Lung’ or ‘Chest’ or ‘Thorax’ or ‘Thoracic’ or ‘Pulmonary’
2. ‘Ultrasound’ or ‘Ultrasonography’ or “Sonography’ or ‘Sonographic’ or ‘Ultrasonic’
3. ‘COVID’ or ‘COVID-19’ or ‘Coronavirus’ or ‘SARS’
4. ‘1’ and ‘2’ and ‘3’
5. Remove duplicates from ‘4’

## Supplementary material 2: Screening and selection tool

The following screening and selection tool was used by two independent reviewers during abstract and full-text review.

| LUS in COVID-19: Screening and Selection Tool |  |  |
| --- | --- | --- |
| Reviewer name: |  | Date: |
| Title: |  |  |
| Author name: | Publication Date: | Journal: |
|  | Inclusion Criteria | Exclusion Criteria |
| Diagnostic test: | ☐ LUS |  |
| Population: | ☐ Age >16<br>☐ Acute presentation of suspected COVID-19 |  |
| Intervention: | ☐ Lung Ultrasound |  |
| Comparator: | ☐ CT<br>☐ PCR<br>☐ Final Diagnosis |  |
| Outcomes: | ☐ Diagnostic Accuracy |  |
| Study design: | ☐ Prospective<br>☐ Retrospective<br>☐ Experimental<br>☐ Observational | ☐ Case reports / series<br>☐ Editorials<br>☐ Recommendations<br>☐ Reviews |
| Overall decision: | ☐ Included | ☐ Excluded |
| Notes: |  |  |

